# CAVIN1-Mediated Endocytosis: A Novel Mechanism Underlying The Interindividual Variability In Drug-Induced Long QT

**DOI:** 10.1101/2023.02.21.23286271

**Authors:** Zeina R. Al Sayed, Céline Pereira, Esthel Pénard, Adeline Mallet, Charlène Jouve, Nihar Masurkar, Gildas Loussouarn, David-Alexandre Trégouët, Jean-Sébastien Hulot

## Abstract

**Background:** Drug-induced QT prolongation (diLQT) is a feared side-effect as exposing susceptible individuals to fatal arrhythmias. The occurrence of diLQT is primarily attributed to unintended drug interactions with cardiac ion channels, notably the hERG channels that generate the repolarizing current (IKr) and thereby regulate the late repolarization phase. There is an important inter-individual susceptibility to develop diLQT which is of unknown origin but can be reproduced in patient-specific iPSC-derived cardiomyocytes (iPS-CMs).

**Objective:** We aimed to investigate the dynamics of hERG channels in response to sotalol and to identify regulators of the susceptibility to developing diLQT.

**Methods:** We measured electrophysiological activity and cellular distribution of hERG channels after hERG blocker treatment in iPS-CMs derived from patients with highest or lowest sensitivity (HS or LS) to sotalol administration *in vivo* (*i*.*e*., based on the measure of the maximal change in QT interval 3 hours after administration). Specific small-interfering RNAs (siRNA) and CAVIN1-T2A-GFP adenovirus were used to manipulate *CAVIN1* expression.

**Results:** While HS and LS iPS-CMs showed similar electrophysiological characteristics at the baseline, the late repolarization phase was prolonged, and I_Kr_ significantly decreased after exposure of HS iPS-CMs to low sotalol concentrations. I_Kr_ reduction was caused by a rapid translocation of hERG channel from the plasma membrane to the cytoskeleton upon sotalol application. This phenomenon was suppressed by blocking active endocytosis using dynasore. *CAVIN1*, essential for caveolae biogenesis, was two-times more expressed in HS iPS-CMs and its knockdown using siRNA decreased their sensitivity to sotalol. *CAVIN1* overexpression in LS iPS-CMs using adenovirus showed reciprocal effects. Mechanistically, we found that treatment with sotalol promoted trafficking of the hERG channel from the plasma membrane to the cytoskeleton through caveolae and in a manner dependent on CAVIN1 expression. *CAVIN1* silencing reduced the number of caveolae at the membrane and abrogated the internalization of hERG channel in sotalol-treated HS iPS-CMs. CAVIN1 also controlled cardiomyocyte responses to other hERG blockers such as E4031, vandetanib, and clarithromycin.

**Conclusions:** Our study identifies unbridled turnover of the potassium channel hERG as a mechanism supporting the inter-individual susceptibility underlying diLQT development and demonstrates how this phenomenon is finely tuned by CAVIN1.

**CLINICAL PERSPECTIVE:** *What is new?:*

- The inter-individual susceptibility underlying diLQT development involves unbridled turnover of cardiac ion channels from the plasma membrane.
- This phenomenon is finely tuned by CAVIN1, a protein that is essential for essential for caveolae biogenesis.
- Treatment with hERG blocker promoted trafficking of the hERG channel from the plasma membrane to the cytoskeleton through caveolae and in a manner dependent on CAVIN1 expression.

*What are the clinical implications?:*

- While congenital long QT is primarily from a genetic origin, the development of drug-induced long QT involves differences in the trafficking machinery of cardiac ion channels.
- The prediction of CAVIN1 expression levels could help preventing drug-induced cardiotoxicity.

## INTRODUCTION

Interindividual variability in drug response is unpredictable and remains largely unexplained. While most patients develop an appropriate drug response in terms of efficacy and safety, some patients treated with cardiac or non-cardiac medications experience adverse side-effects that can be life-threatening. The development of drug-induced QT prolongation (diLQT) and the consequent fatal arrhythmias (i.e., Torsade de Pointes) in some patients is one of the most striking examples of cardiotoxicity^1^. The occurrence of diLQT is primarily related to drug interactions with cardiac ion channels, notably the hERG channels that regulate the late repolarization phase through the rapid component of the cardiac delayed rectifier potassium current (named I_Kr_). This channel constitutes a common unintended target of drugs from almost every therapeutic class, including antiarrhythmic, anti-psychotic, antibiotic, antihistamine, and anticancer agents^1, 2^. Why some patients are more susceptible than others and display excessive inhibition of hERG channels in response to the same pharmacological stimulation is, however, unclear^3^.

As mutations in *KCNH2* (the gene encoding hERG) are detected in patients with congenital LQT syndrome, it has been proposed the existence of genetic determinism to diLQT^4-7^. However, genome-wide screening studies in large cohorts of patients with diLQT have globally ruled out the essential contribution of variants of *KCNH2* or other genes involved in congenital LQT ^8, 9^.

We and others have previously reported that the predilection to diLQTS as observed *in vivo* could be reproduced *in vitro* using induced pluripotent stem cell derived-cardiomyocytes (iPS-CMs)^10, 11^. These studies suggest the *in vitro* reproduction of a peculiar mechanism that supports the interindividual variability in the susceptibility to develop diLQTS. Here, we exploited iPS-CMs derived from individuals with differential sensitivity to sotalol^10^, a class III antiarrhythmic drug and an unintentional hERG blocker, to investigate any common molecular mechanism promoting high sensitivity to hERG blockers. By comparing the transcriptomic profiles of iPS-CMs for individuals with extreme response to sotalol, we previously identified five differentially expressed genes (*CAMKV, DLG2, KCNE4, HTR2C*, and *CAVIN1*) that were mainly related to the regulation of cardiac repolarization^10^. Here, our data demonstrate that *CAVIN1*, an essential gene for caveolae biogenesis, mediates diLQT. High expression levels of this gene drove hERG internalization upon drug exposure, leading to I_Kr_ reduction and the consequent prolongation in the repolarization phase in iPS-CMs derived from individuals presenting with sotalol-induced LQT.

## MATERIALS AND METHODS

Details on the procedures are provided in the online-only data supplement. The data and material that support the findings of this study are available from the corresponding author upon reasonable request.

### iPSC Maintenance in Cultures

Skin biopsies from 10 individuals with the highest QT interval prolongation (HS) and 10 individuals with the lowest change in QT (LS) after sotalol administration were reprogrammed into iPSCs, as previously reported^10^. In the current study, we selected three cell lines for HS and three cell lines for LS groups that showed concordant *in vivo* and *in vitro* response to sotalol, and with respect to gender (2 females and 1 male in each group)^10^. iPSC were expanded on stem cell-qualified Matrigel-coated (Corning) plates with mTeSR1 (StemCell technologies). At 80% confluency, cells were passaged using ReleSR™ passaging reagent (StemCell Technologies).

### Differentiation of iPSCs into Cardiomyocytes

At 90% confluency, iPSCs were placed in Roswell Park Memorial Institute (RPMI)-1640 medium (Life Technologies) supplemented with B27 without insulin (Life Technologies) and 6 μM CHIR-99021 (Abcam). After 48 h, the medium was changed to RPMI-1640-B27 without insulin. The following day, the medium was replaced with RPMI-1640-B27 without insulin supplemented with 5 μM IWR-1 (Sigma). By day 5, cells were cultured in RPMI-1640-B27 without insulin and then switched into RPMI1640-B27 with insulin after 48 h. On day 11, beating iPS-CMs were subjected to glucose starvation in RPMI-1640-B27 without glucose (Life Technologies) for 3 days. Cells were then dissociated using 0.05% trypsin (Life Technologies) for 5 min and seeded at a 1.2 × 10^6^ cells/well density. The following day, the medium was switched to RPMI-B27 with insulin for 24h, after which the cells were subjected to a second round of glucose starvation for 3 days. By day 16, cells were cultured in RPMI1640-B27 with insulin and the medium was changed every two days. All experiments were performed around day 30 of differentiation.

### Immunofluorescence

iPSCs and iPS-CMs at day 30 of differentiation were plated on Matrigel-coated coverslips. Cells were then fixed in 4% paraformaldehyde for 15 min, permeabilized with Triton X-100, blocked with 2% bovine serum albumin (BSA), and incubated overnight with appropriate primary antibodies (Table S1). The cells were then probed with appropriate conjugated secondary antibodies and stained with 4′,6-diamidino-2-phenylindole (DAPI). Immunostaining was examined using a Leica SPE confocal system.

### Electrophysiological Assessment

#### Field potential (FP) recording

FP recording was conducted using six-well MEA arrays (60-6 well MEA 200/30 iR-Ti-rcr, Multichannel Systems) or one-well MEA arrays (MEA 200/30 iR-Ti-rcr, Multichannel Systems). Four days prior to recording, iPS-CMs were dissociated using Enzyme T (Miltenyi Biotec) for 10 min at 37°C, and seeded on the electrode array coated with Matrigel at a density of 200,000 cells/3 μL medium. After 4 h, RPMI-1640-B27 insulin medium was added to the system; the medium was changed every other day. Each drug sotalol (Selleckchem), E4031 (StressMarq), clarithromycin (Selleckchem), and vandetanib (Selleckchem), was prepared at different concentrations in RPMI medium supplemented with B27 and insulin and directly added to wells during recording at 37°C. Methyl-β-cyclodextrin (MβCD; Sigma) and Dynasore (Sigma) were added respectively 15 min and 60 min before the recordings and pharmacological stimulations. FP was recorded using Cardio 2D software (Multichannel Systems) for 3 min for each condition preceded by 3 min of stabilization following drug treatment. Raw MEA data were analyzed using Cardio 2D+ (Multichannel Systems). FP values were averaged from 2 min recording by the software. Field potential duration (FPD) was then determined and normalized to peak-to-peak duration using Frederica’s formula as follows: FPDc = FPD/^3^√(peak-to-peak duration).

#### Patch clamp

On days 21-24 of differentiation, iPS-CMs were dispersed as single cells using Enzyme T (Miltenyi Biotec) for 10 min at 37°C and plated at 60000 cells/35-mm Petri dish. After 10-14 days from seeding, patch-clamp recordings were performed using an Axopatch 200B amplifier controlled by Axon pClamp 11 software through an A/D converter (Digidata 1440B). All recordings were conducted at 37°C. In current clamp, stimulation was performed using custom-made software running on RT-Linux and an A/D converter (National Instrument PCI-6221) connected to the current command of the amplifier^12^. Data were collected from at least three independent differentiation from each group and analyzed using Clampfit 11 software (Molecular Devices).

#### Current clamp

Action potential (AP) was acquired in a perforated-patch configuration using 0.22 mM amphotericin-B (Sigma). Cells were constantly perfused using Tyrode’s solution containing 140 mM sodium chloride (NaCl), 4 mM potassium chloride (KCl), 1 mM calcium chloride (CaCl_2_), 0.5 mM magnesium chloride (MgCl_2_), 10 glucose, 10 HEPES (pH 7.4 adjusted with sodium hydroxide [NaOH]). Patch pipettes (tip resistance: 3-5 MΩ) were filled with a solution containing 125 mM K-gluconate, 20 mM KCl, 5 mM NaCl, 5 mM HEPES (pH 7.2 adjusted with potassium hydroxide [KOH]), and amphotericin-B. After 2 min of spontaneous AP recording, cells were paced with 1 ms 30-50 pA/pF stimulation pulse at 500 and 700 ms of cycle length. AP classification into ventricular type was determined from the evaluation of the ratio (APD30-APD40)/(APD70-APD80) > 1.45, which reflects the presence of a plateau phase. Seven consecutive APs were superposed and averaged. Sotalol solubilized in dimethyl sulfoxide (DMSO) was used at 30, 50, and 100 μM concentrations in a solution containing 140 mM NaCl, 4 mM KCl, 1 mM CaCl_2_, mM 0.5 MgCl_2_, 30 mM mannitol, 10 mM HEPES; (pH 7.4 adjusted with NaOH).

#### I_Kr_ recordings in iPS-CMs

I_Kr_ recordings were performed in the ruptured whole-cell configuration. Voltage clamp protocols are depicted in the corresponding figures. Borosilicate pipettes with 3-4 MΩ tip resistance were used for recordings at 1 KHz low-pass filter and filled with a solution containing 140 mM KCl, 5 mM EGTA, 4 mM MgATP, 1 mM HEPES, and 1 mM MgCl_2_. Current densities were calculated by dividing the current value by membrane capacitance (Cm). A steady-state activation curve was fitted using Boltzmann equation.

#### Reverse-Transcriptase Quantitative Polymerase Chain Reaction (RT-qPCR)

Total RNA (tRNA) was isolated using NucleoSpin RNA kit (MACHEREY-NAGEL) and reverse transcribed using SuperScript IV VILO Master Mix (Thermo Fisher Scientific). qPCR was carried out with SYBR Select Master Mix (Applied Biosystems) using the primers listed in Table S2. Gene expression was determined according to the ΔΔCt method following normalization to the expression of the housekeeping gene *RPL32*.

### Protein Extraction

#### Total protein lysate

Total protein was extracted using a lysis buffer containing 1% TritonX-100; 100 mM NaCl, 50 mM Tris-HCl, 1 mM EGTA, 1 mM sodium orthovanadate (Na_3_VO_4_); 50 sodium fluoride (NaF), phosphatase inhibitor cocktails 2 and 3 (P0044 and P5726, Sigma-Aldrich), and protease inhibitors cocktail (P8340, Sigma-Aldrich). Extracted samples were sonicated and centrifuged at 15,000 ×*g* for 15 min at 4°C. Protein quantification was performed using Pierce™ BCA Protein Assay Kit (Thermo Fisher).

#### Cell fraction isolation

Proteins from the cytoplasm, membrane, nucleus, and cytoskeleton compartments were isolated from at least 5 million cells per condition using Qproteome Cell Compartment Kit (Qiagen), as per the manufacturer’s instructions. Extracted proteins were precipitated using acetone, solubilized in the lysis buffer mentioned before, and quantified using Pierce™ BCA Protein Assay Kit (Thermo Fisher). Fractioning was validated by revealing a specific protein from each cell compartment.

#### Viral Infection

On day 21-23 of differentiation, cardiomyocytes were infected with an adenovirus (AdV) carrying cytomegalovirus (CMV) promoter driving CAVIN1 plus a T2A separating EGFP (VB190717-1058jag) and AdV carrying CMV driving GFP (VB150925-10024) only as a control (Vector Builder) at a multiplicity of infection (MOI) of 50 for overnight. The medium was replaced with fresh RPMI-1640-B27 with insulin supplemented with 5% fetal bovine serum (FBS). Seven days following the infection, iPS-CMs were used for subsequent experiments.

#### CAVIN1 Silencing

Small-interfering RNA (siRNA) targeting *CAVIN1* (siCAVIN1; s49507), non-targeting siRNA (siNeg; 4390843), and GAPDH silencer (siGAPDH; 4390849) were purchased from Thermo Fisher Scientific. GAPDH siRNA was used to determine the optimal siRNA quantity that allowed maximum knockdown, and 200 nM siRNA was sufficient to induce 70% reduction in the RNA expression of GAPDH and was used for the following transfection. On day 24-25 of differentiation, transfection with either siCAVIN1 or siNeg was performed using Lipofectamine™ RNAiMAX Transfection Reagent (Thermo Fisher Scientific) as per the manufacturer’s instructions. Cells were used after 5 days of transfection.

#### Statistical Analysis

The number of samples (*n*) used in each experiment is recorded in the text and figure legends. All experiments were performed independently at least twice. Results are expressed as mean ± standard error of means (SEM). Comparisons were made using Mann-Whitney U test for comparison between two groups, Kruskal-Wallis test or two-way analysis of variance (ANOVA) for comparison between > two groups. Bonferroni post-hoc test was used for repeated measures. A value of p < 0.05 was considered statistically significant. Statistical analyses were performed with GraphPad Prism software (Version 9).

## RESULTS

### Patient-Specific iPS-CMs Recapitulate Inter-Individual Variability in Sotalol Sensitivity

We used 6 lines from our previously developed library of patient-specific iPSC^10^, 3 of them (2 females, 1 male) derived from individuals with the highest sensitivity (HS) and 3 (2 females, 1 male) with lowest sensitivity (LS) to sotalol (*i*.*e*., based on the measure of the maximal change in QT interval 3 h after sotalol administration). Expression of pluripotency markers was validated in all clones (Figures S1A-B), and cardiac differentiation efficiency was assessed by immunostaining for cardiac markers (troponin T and α-actinin) (Figure S1C) and flow cytometry. Results revealed an average of 98.9% troponin T-positive cells in both groups (Figure S1D). A predominance of the ventricular-like type (∼85%) was observed after analyzing action potential recordings (Figure S1E). The ratio of the ventricular (MYL2) and atrial (MYL7) myosin light chain RNA expression was similar between HS and LS iPS-CMs (Figure S1F).

We then compared the electrophysiological parameters of HS and LS iPS-CMs (n = 3 different clones for each group) using the patch-clamp technique at the single-cell level and MEA recordings at the tissue level. These iPS-CMs reproduced the clinical phenotype of patients as they exhibited similar electrophysiological parameters at baseline and more pronounced prolongation of the repolarization durations in response to sotalol in HS iPS-CMs (Figures 1A-1F, Figure S2 and Table S3). APD90 and APD50 were significantly higher in HS than in LS iPS-CMs at 30 and 50 μM sotalol concentrations (Figure 1D, Figure S2C and Table S3). These results were reproduced at the tissue level (Figures 1G and Table S3). While the late phase of repolarization was affected, 30 and 50 μM sotalol neither changed the early stage of repolarization (APD30) (Figure 1C) nor the resting membrane potential (Figure 1B) and the depolarization phase (velocity, amplitude, and overshoot) (Figures 1E-1F, Figure S2D and Table S3). This observation revealed a principal effect of sotalol on late repolarization currents in iPS-CMs derived from patients with high sensitivity to sotalol *in vivo*.

**Figure 1:**
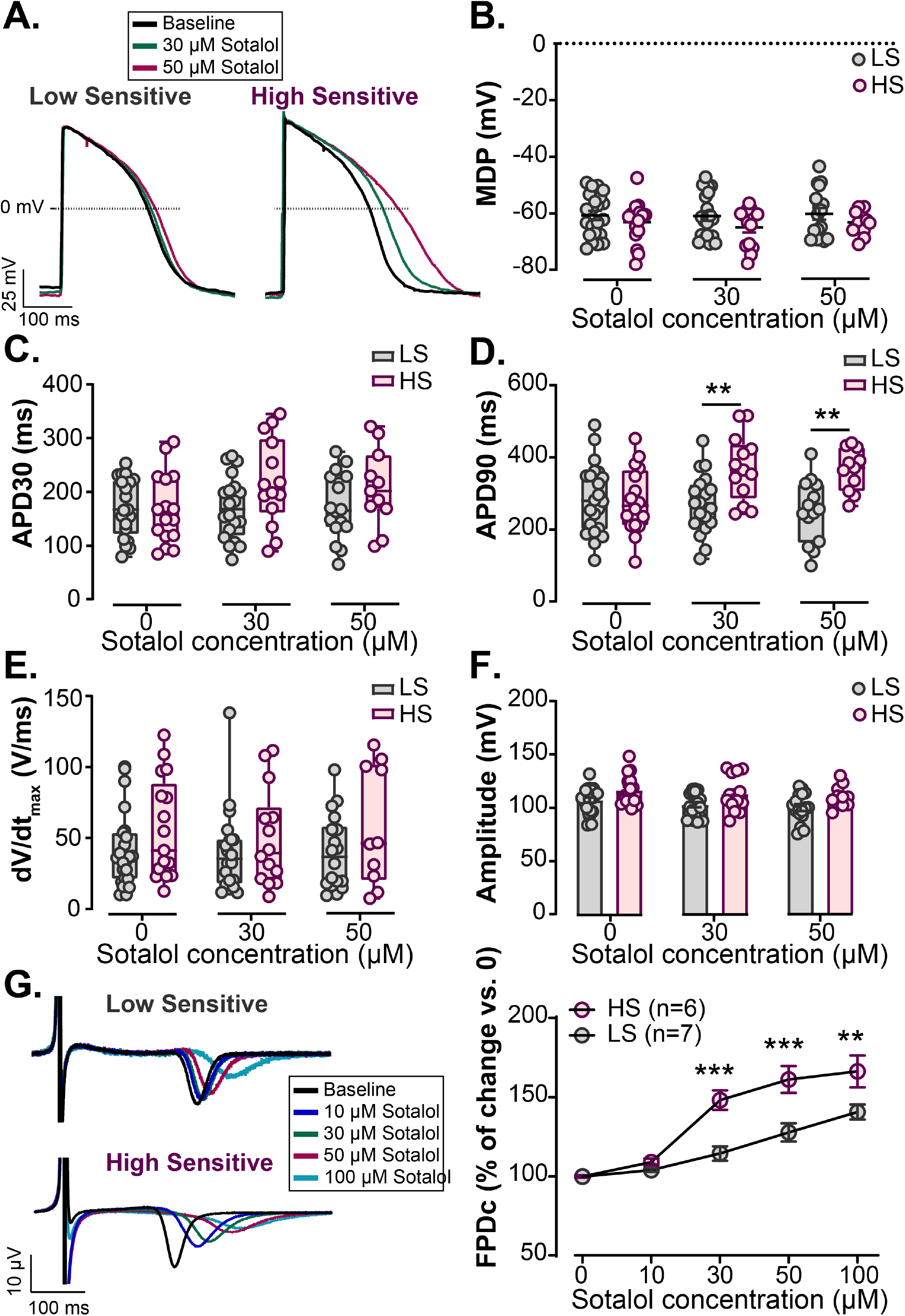
HS iPS-CMs, but not LS iPS-CMs exhibit a prolongation in the late repolarization phase after sotalol application. A. Aligned APs comparing the baseline with 30 and 50 μM sotalol treatments in the low and high-sensitive iPS-CMs. B. Membrane diastolic potential (MDP) measured at baseline (0) and after sotalol application (30 and 50 μM) in LS and HS iPS-CMs. C. Box whisker blot of APD at 30% repolarization (early phase) analyzed from LS and HS iPS-CMs paced at 1.5 Hz before and after applying 30 and 50 μM sotalol. D. Box whisker blot of APD at 90% repolarization (late phase) analyzed from LS and HS iPS-CMs paced at 1.5 Hz treated or not with 30 and 50 μM sotalol. **p < 0.01 *vs*. LS iPS-CMs (two-way ANOVA with Bonferroni post-hoc test). E. Maximum upstroke velocity dV/dt_max_ calculated in LS and HS iPS-CMs paced at 1.5 Hz after treatment with different concentrations of sotalol. F. Upstroke amplitude analyzed from the action potential recorded in HS and LS iPS-CMs treated with different concentrations of sotalol and paced at 1.5 Hz. G. Right: Aligned FPs showing the impact of different concentrations of sotalol on LS and HS iPS-CMs. Left: Percentage of FPDc change normalized *vs*. baseline following the application of sotalol. ** and ***p < 0.01 and p < 0.001 *vs*. LS iPS-CMs, respectively (two-way ANOVA with Bonferroni post-hoc test).

### Sotalol Application Changes hERG Distribution within HS iPS-CMs

The expression levels of *KCNH2* and other central ion channel genes (*KCNQ1, SCN5A, CACAN1C*, and *KCNE2*) implicated in congenital long QT syndrome were not different between HS and LS iPS-CMs (Figure 2A and Figure S3A). At the protein level, the mature glycosylated hERG (molecular weight (MW) of 150 KDa) expressed at the membrane and the non glycosylated form (MW of 135 KDa) were similarly expressed between both groups (Figure 2B). At the functional level, the density and activation properties of I_Kr_, which is the current generated by hERG channels, were not different between HS and LS iPS-CMs before any pharmacological stimulation (Figure 2C and Figures S3B-E). However, I_Kr_ maximum tail current density was significantly and profoundly decreased in HS iPS-CMs after treatment with 30 and 50 μM sotalol but remained unchanged in LS iPS-CMs (Figure 2C). Overall, these results indicate a peculiar reactivity of hERG channels in HS iPS-CMs to sotalol. This phenomenon was consistent with a decrease in I_Kr_ that correlates with the higher prolongation of repolarization after sotalol intake, as observed in patients with a predisposition to diLQT.

**Figure 2:**
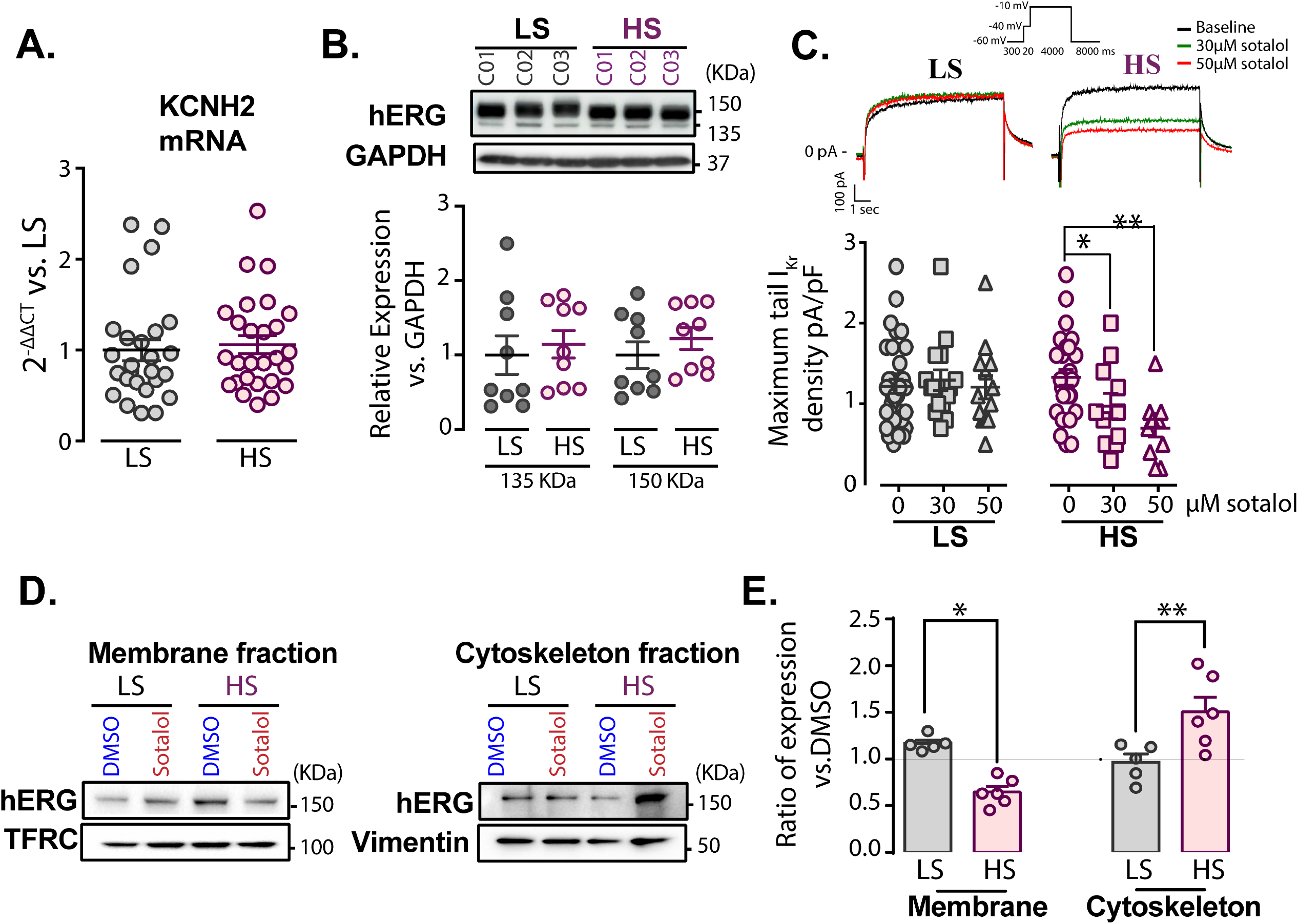
Changes in the repolarization phase of HS iPS-CMs after sotalol application are due to different hERG distribution within the cell. A. KCNH2 RNA expression level at day 30 of differentiation measured by SYBR green in LS (n = 26) and HS iPS-CMs (n = 27). Cts were normalized to RPL32 and the ratio *vs*. LS (2^−ΔΔCt^) was then calculated. B. Representative immunoblot of hERG encoded by KCNH2 from the total protein lysate at day 30 of differentiation of three different clones of LS and HS iPS-CMs and the ratio of membranous hERG of 150 KDa and cytosolic immature hERG of 135 KDa expression normalized to GAPDH. C. Top: Superimposed traces for I_Kr_ before (black) and after (colored) the application of 30 and 50 μM sotalol in LS and HS iPS-CMs. Bottom: Maximum tail I_Kr_ density without or with application of different concentrations of sotalol. * and **p < 0.05 and p < 0.01 *vs* before sotalol application, respectively (Kruskal-Wallis test). D. Left: Representative immunoblot of hERG expression in different LS and HS iPS-CM fractions treated with either 0.1% DMSO or 50 μM sotalol for 10 min. TFRC and vimentin were used to normalize the plasma membrane and cytoskeleton fraction, respectively. Right: Ratio of hERG (150 KDa) expression after treatment with 50 μM sotalol *versus* DMSO in LS and HS iPS-CMs. * and **p < 0.05 and p < 0.01 *vs*. LS iPS-CMs, respectively (one-way ANOVA).

Total hERG (glycosylated and non-glycosylated) levels were unaffected by sotalol treatment (Figures S4A-D). To test whether the decrease in I_Kr_ is associated with altered hERG expression trafficking, we traced hERG distribution in different cellular fractions (Figure S4E) in HS and LS iPS-CMs treated with vehicle (DMSO) or sotalol. hERG expression was significantly lower in the cell membrane but increased in the cytoskeleton fraction of HS iPS-CMs in response to sotalol treatment (Figure 2D-E); no significant change in hERG expression was detected in the cytoplasmic fraction (Figure S4F). In contrast, the subcellular expression of hERG was unchanged in all cellular compartments of LS iPS-CMs after sotalol treatment (Figure 2D-E). Considering the intact hERG levels in the cytoplasmic fraction, we decided to exclude this fraction from further investigations. Overall, our results are in line with the original mechanism where higher susceptibility to develop abnormal cardiac repolarization in response to an offending drug is supported by the drug-induced internalization of targeted channels from the membrane to the cytoskeleton.

### CAVIN1 Expression Level Determines the Sensitivity to Sotalol

Among the five differentially expressed genes (*CAMKV, DLG2, KCNE4, HTR2C*, and *CAVIN1*) previously identified^10^, we confirmed the higher mRNA expression of *CAVIN1* in HS than in LS iPS-CMs (Figure 3A); however, no significant differences in the expression of other candidates (Figure S5). The protein expression of CAVIN1 (also called PTRF) was also two-fold higher in HS iPS-CMs than in LS iPS-CMs (Figure 3B). In line with our electrophysiological observations, CAVIN1 appeared as an appealing candidate previously reported to regulate the remodeling and trafficking of caveolae, a type of cell membrane structure containing cardiac ion channels^13^. Confocal microscopy images confirmed the sub-membranous localization of CAVIN1 in alpha-actinin-positive LS and HS iPS-CMs (Figure 3C).

**Figure 3:**
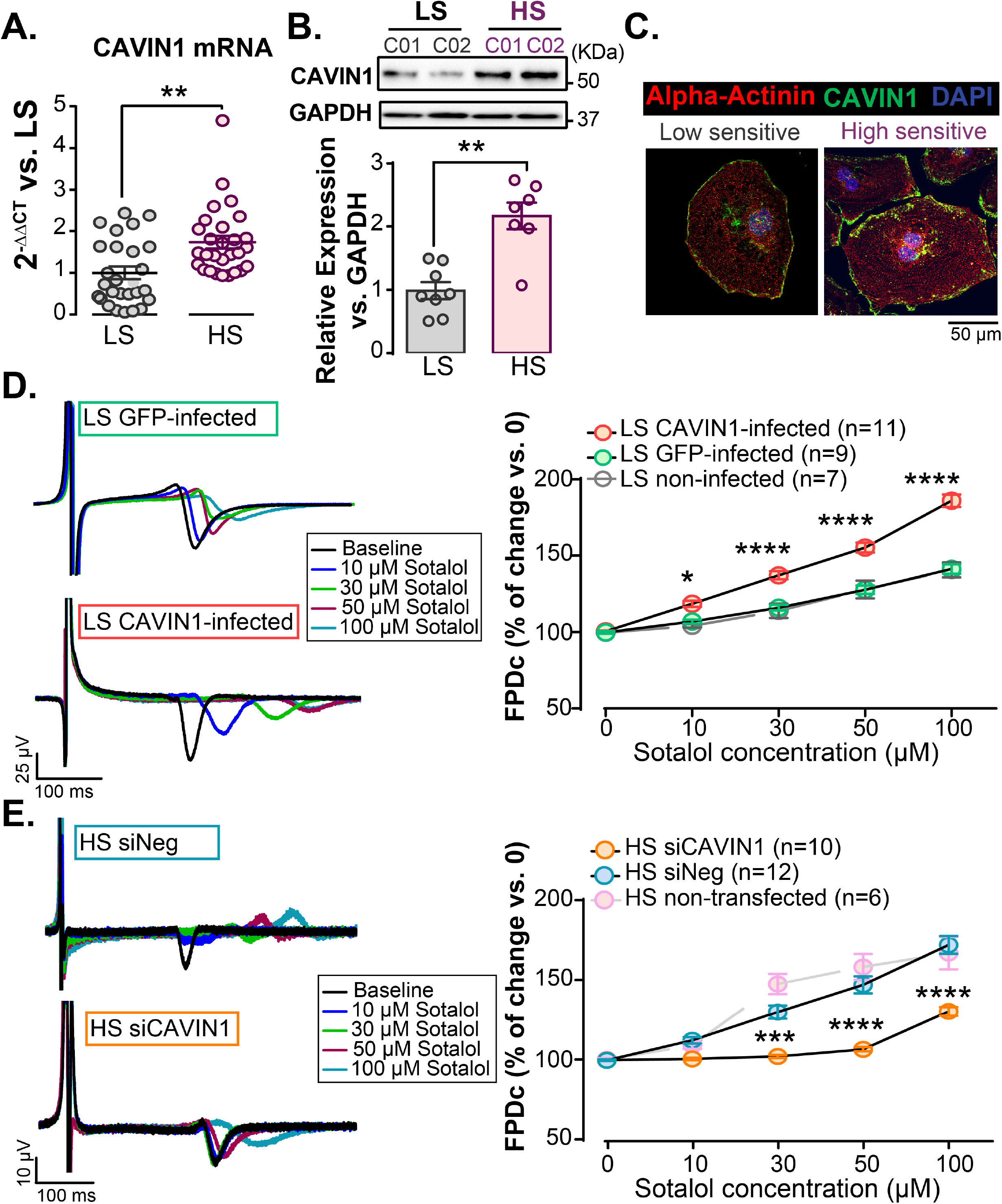
Modulation of CAVIN1 expression in HS and LS iPS-CMs is sufficient to reverse the changes in the repolarization duration in response to sotalol. A. Comparison of the transcriptomic expression of CAVIN1 between LS and HS iPS-CMs. RT-qPCR data are presented as relative fold change with reference to LS iPS-CMs. **: p < 0.01 *vs*. LS iPS-CMs (Mann-Whitney test). B. Top: Representative immunoblot illustrating CAVIN1 expression in LS and HS iPS-CMs. Bottom: Expression level of CAVIN1 in total protein lysate extracted from LS (n = 8) and HS iPS-CMs (n = 7) normalized to GAPDH. *: p < 0.05 *vs*. LS iPS-CMs (Mann-Whitney test). C. Representative confocal images for immunostainings of CAVIN1 (green) and the cardiac marker α-actinin (red) in LS and HS iPS-CMs. Nuclei are stained in blue using DAPI. D. Left: Representative aligned field potential (FP) recorded from LS iPS-CMs infected with either GFP or CAVIN1 adenovirus before and after applying different concentrations of sotalol. Right: Percentage of change as compared to baseline of corrected FPD (FPDc) measured in LS iPS-CMs infected with either GFP- (green) or CAVIN1-T2A-GFP-adenovirus (red) in response to increasing concentrations of sotalol. FPDc values from non-infected LS iPS-CMs are presented in grey. * and *** p < 0.05 and p < 0.001 *vs*. GFP-infected LS iPS-CMs, respectively (two-way ANOVA with Bonferroni post-hoc test). GFP-infected: n = 9, CAVIN1-infected: n = 11, and non-infected LS iPS-CMs: n = 7. E. Left: Superimposed FP recorded from HS iPS-CMs 5 days following the transfection with either siNeg or siCAVIN1 before and after application of increasing concentrations of sotalol. Right: Percentage of change compared to baseline FPDc measured in HS iPS-CMs transfected with siNeg or siCAVIN1. FPDc values from non-transfected HS iPS-CMs are plotted in pink. *** and **** p < 0.001 and p < 0.0001 *vs*. siNeg-transfected HS iPS-CMs, respectively (two-way ANOVA with Bonferroni post-hoc test). SiNeg: n = 12, siCAVIN1: n = 10, and non-transfected: n = 6.

To further investigate the role of CAVIN1 in the repolarization response to sotalol, we performed experiments that reversed CAVIN1 expression in HS and LS groups. First, we over-expressed CAVIN1 in LS iPS-CMs using an adenovirus harboring *CAVIN1*. Seven days after infection, the protein expression of CAVIN1 increased by around 10-fold as compared to that in LS iPS-CMs infected with the control GFP-adenovirus (Figure S6A). MEA recordings showed a dramatic change in the response of LS iPS-CMs overexpressing CAVIN1 to sotalol, as only 10 μM sotalol was sufficient to induce a significant prolongation in the repolarization duration as compared to controls (Figure 3D). The beating rate changed at 50 and 100 μM sotalol concentrations (Figure S6B). Then, we knocked down *CAVIN1* expression in HS iPS-CMs after verifying 200nM of siRNA as the optimal quantity to induce maximum RNA reduction (Figure S6C). Five days after *CAVIN1*-siRNA transfection, CAVIN1 expression decreased by 84% at the RNA level (Figure S6D) and 62% at the protein level (Figure S6E). This reduction in the CAVIN1 expression in HS iPS-CMs changed their reaction to sotalol, as evident from a significantly lower reactivity to 30, 50, and 100 μM concentrations of sotalol (Figure 3E) without any change in the beating rate as compared to siNeg-transfected cells (Figure S6F). Hence, CAVIN1 expression modulation was sufficient to strongly reverse the specific phenotypes observed in our patient-specific iPSC library, indicating CAVIN1 as a significant driver of the individual susceptibility to the drug response.

### Disruption of Caveolae Endocytosis Limits the Impact of CAVIN1 on Sotalol Sensitivity

In order to further confirm the involvement of caveolae, we first identified the presence of caveolae at the plasma membrane of iPSC-CMs using transmission electron microscopy (TEM), and observed that the caveolae density was similar in LS and HS iPS-CMs (Figure S7A). However, in line with the suspected involvement of CAVIN1 in caveolae biogenesis, we observed that the caveolae density at the plasma membrane of HS iPS-CM was significantly decreased after treatment with CAVIN1 siRNA as compared to control (Figure 4A). Concordantly, the expression of caveolin-3 (CAV3) was increased in LS iPS-CMs overexpressing CAVIN1 (Figure S7B). The expression of caveolin1 (CAV1), another essential element in caveolae biogenesis besides CAVIN1, was detected in the cytoskeleton fraction of iPS-CMs (Figure 4B), consistent with its role in anchoring caveolae to the cytoskeleton^14^. CAV1 expression increased in LS iPS-CMs infected with CAVIN1 as compared to that in controls (Figure 4B), thereby confirming the relationship between CAVIN1 expression and caveolae formation in iPS-CMs. In contrary to CAVIN1, *CAV1* and *CAV3* transcript levels were similar between LS and HS iPS CMs at the baseline (Figure S7C).

**Figure 4:**
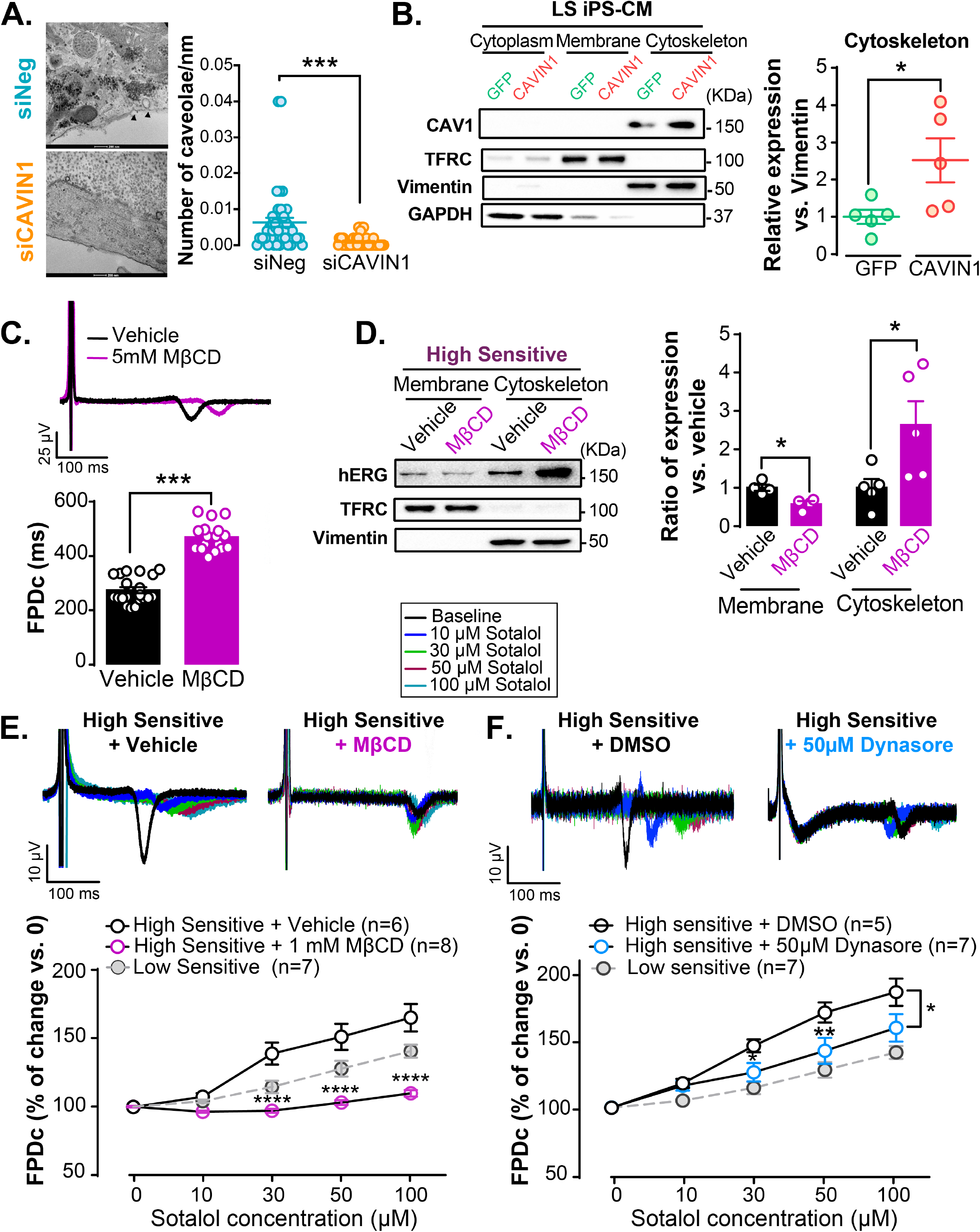
Disruption of caveolae limits CAVIN1 impact on the repolarization phase. A. Left: TEM images showing surface caveolae (indicated with black arrows, counted as omega-shaped membrane forms) in HS iPS-CM treated with scrambled (siNeg) and siRNA against CAVIN1 (siCAVIN1). Right: Quantification of the caveolae density on the cell membrane in siNeg (44 images) and siCAVIN1 (52 images). ***: p< 0.001 Mann-Whitney test. B. Left: Immunoblot displaying CAV1 and CAVIN1 protein expression in different cell compartments and the housekeeping genes for each fraction (TFRC, vimentin, and GAPDH for the membrane, cytoskeleton, and cytoplasmic fraction, respectively). Right: Ratio of CAV1 expression in the cytoskeleton fraction of LS iPS-CMs at day 7 of infection with GFP and CAVIN1-Adv. *: p < 0.05 *vs*. LS iPS-CMs (Mann-Whitney test). C. Top: Representative recordings of FP of HS iPS-CMs treated with either 1 mM of MβCD for 15 min at 37°C or vehicle (water). Bottom: Corrected FPD (FPDc) is compared. *** p < 0.001 *vs*. vehicle (Mann-Whitney test). D. Left: Representative immunoblot of membranous and cytoskeletal hERG isolated from HS iPS-CMs treated with 5 mM MβCD for 15 min at 37°C or vehicle. TFRC and vimentin were used as loading references for the plasma membrane and cytoskeleton, respectively. Right: hERG expression was normalized to each loading reference. * p < 0.05 *vs*. HS iPS-CMs treated with vehicle (Kruskal-Wallis test). n = 5. E. Top: Representative superimposed FPs recorded from HS iPS-CMs treated with either 1 mM MβCD or vehicle for 15 min at 37°C. The 3 HS-iPS-CMs lines were then treated with increasing concentrations of sotalol (with at least 2 independent differentiations per conditions). Bottom: Percentage of change as compared to baseline was calculated and plotted. **** p < 0.0001 *vs*. HS iPS-CMs treated with vehicle. (two-way ANOVA with Bonferroni post-hoc test). F. Top: Aligned FPs recorded from HS iPS-CMs treated with either 0.1% DMSO or 50 μM Dynasore for 1 hour at 37°C. Different concentrations of sotalol were then applied. Bottom: Percentage of change as compared to baseline was calculated and plotted. * p < 0.05 and ** p < 0.01 *vs*. HS iPS-CM treated with DMSO (two-way ANOVA with Bonferroni post-hoc test).

To evaluate the presence of hERG channels in caveolae, we then treated cells with MβCD, a detergent that removes cholesterol from the membrane and, thus, remove caveolae. The application of 1 mM MβCD significantly prolonged the repolarization phase before any pharmaceutical stimulation, indicative of the elimination of a repolarizing channel from caveolae by MβCD (Figure 4C). To assess whether this channel is hERG, we isolated proteins from different cellular compartments after treatment with either 5 mM MβCD or vehicle (water). Immunoblots revealed the decrease in hERG expression in the membranous fraction and its accumulation in the cytoskeleton (Figure 4D). Thus, hERG can be found in the cholesterol-enriched fraction of the membrane. The sensitivity to sotalol was also significantly affected by the removal of caveolae by MβCD. Treatment of HS iPS-CMs with 1 mM MβCD for 15 min strongly reduced the prolongation of repolarization duration upon sotalol application (Figure 4E). Similarly, MβCD-mediated caveolae disruption in LS iPS-CMs overexpressing CAVIN1 significantly limited the prolongation in FPDc at different sotalol concentrations (Figure S8). We further assess the dependence of the process on active endocytosis by using Dynasore, a molecule that blocks dynamin, the protein responsible for caveolar scission from the plasma membrane^15^. Treating the HS iPSC-CM with 50 μM Dynasore for 1 hour prior to electrophysiological recordings, reduced the exacerbated response of the HS iPSC-CMs to sotalol, to a level similar to the one observed with LS iPSC-CMs. Overall, these data suggest a link between CAVIN1, caveolae trafficking, active endocytosis and the increased sensitivity to sotalol.

### CAVIN1 Enhances hERG Channels Turnover in HS iPS-CMs After Sotalol Treatment

We further investigated the expression of different forms of hERG channel (the predominant isoform in adult heart hERG1a 150KDa, hERG1b 120KDa, and degraded hERG 70KDa) in response to sotalol. We first observed that the decrease in the mature hERG1a channel expression in the membrane compartment after sotalol treatment (Figure 2D) was associated with a significant increase in the fragmented form of hERG (Figure 5A) in the cytoskeleton fraction of HS iPS-CMs as compared to that in LS iPS-CMs (Figure 5A). To assess the possibility of hERG trafficking back to the membrane, we compared the recruitment of RAB11, a marker of recycling endosomes, which was mainly detected in the membrane fraction and was more expressed in HS iPS-CMs treated with sotalol than those treated with DMSO. However, in LS iPS-CMs, RAB11 expression level was similar after treatment with sotalol or DMSO (Figure 5B). Overall, sotalol treatment of HS iPS-CMs induced hERG endocytosis into vesicles where some were fragmented, and others were recycled back to the surface.

**Figure 5:**
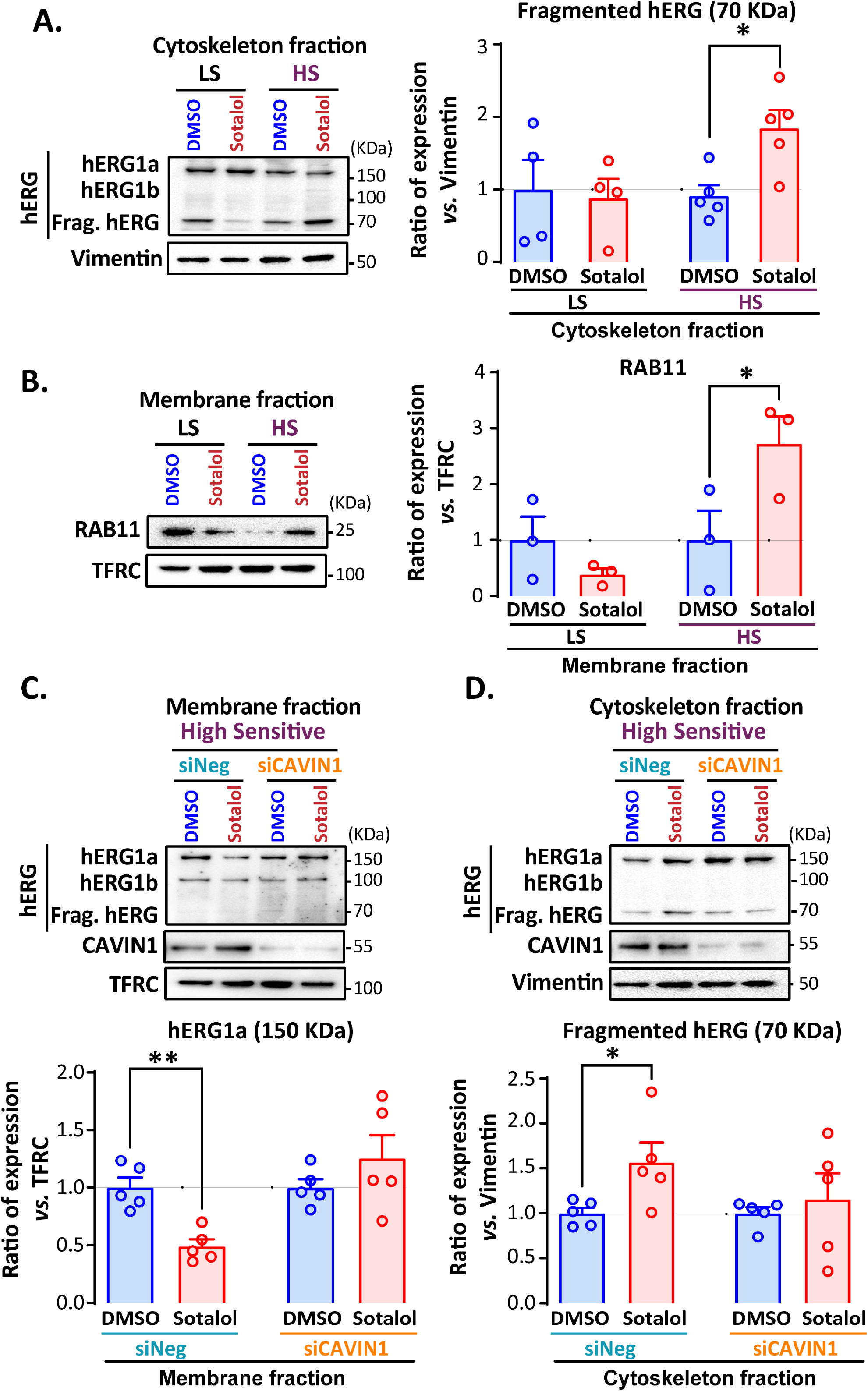
CAVIN1 increases hERG endocytosis upon sotalol treatment in HS iPS-CMs. A. Left: Representative immunoblot of hERG at different molecular weights (hERG1a, hERG1b, and fragmented hERG at 150, 110, and 70 KDa, respectively) in LS and HS iPS-CM cytoskeletal fraction treated with either 0.1% DMSO or 50 μM of sotalol for 10 min. Vimentin was used as a loading reference for the cytoskeleton fraction. Right: Ratio of fragmented hERG (70 KDa) after treatment with sotalol *versus* DMSO in LS (n = 4) and HS iPS-CMs (n = 5) is plotted. * p < 0.05 *vs*. LS iPS-CM, respectively (Kruskal-Wallis test). B. Left: Representative immunoblot of RAB11, CAVIN1, and TFRC in the membranous fraction of LS and HS iPS-CMs treated with sotalol or DMSO. RAB11 bands were normalized to TFRC. Right: * p < 0.05 *vs*. treatment with DMSO (Kruskal-Wallis test). n = 3. C. Top: Representative immunoblot showing hERG and CAVIN1 as well as the loading reference of the membrane fraction (TFRC) in HS iPS-CMs transfected with either siNeg or siCAVIN1 and then exposed to DMSO or sotalol. Bottom: Quantification of mature hERG1a (150 KDa) expression at the plasma membrane normalized to TFRC and ratio compared to DMSO in HS iPS-CMs transfected with CAVIN1 siRNA or negative siRNA. ** p < 0.01 *vs*. treatment with DMSO (Kruskal-Wallis test). n = 4. D. Top: Representative immunoblot of hERG, CAVIN1, and vimentin in the cytoskeleton of HS iPS-CMs transfected with either siNeg or siCAVIN1 and then treated with DMSO or sotalol. Bottom: Fragmented form of hERG (70 KDa) in the cytoskeleton normalized to vimentin and ratio as compared to DMSO in HS iPS-CMs transfected with negative siRNA. * p < 0.05 *vs*. treatment with DMSO (Kruskal-Wallis test). n = 4.

We finally tested whether this mechanism is CAVIN-1 dependent. We analyzed hERG channel endocytosis in response to sotalol in HS iPS-CMs treated with siRNA against CAVIN1 or control (Figure S9). The process involving the reduction in the mature hERG1a channel (150 KDa) at the cell membrane and the accumulation of the fragmented form of hERG in the cytoskeleton of HS iPS-CMs after sotalol treatment was not observed anymore after CAVIN1 silencing as compared to control (Figure 5C-D). These results further confirm that CAVIN1 contributes to the increased ratio of cytoskeleton fraction *versus* membrane fraction of hERG channels in response to sotalol treatment.

### CAVIN1-dependent hERG Channels Respond to other hERG Blockers

We finally measured the response of our patient-specific iPS-CMs to other drugs targeting hERG, including E4031, a known blocker of hERG channel, vandetanib, an anticancer drug with medium risk of Torsade de Pointes, and clarithromycin, an antibiotic with a low risk of Torsade de Pointes. In comparison with LS iPS-CMs, HS iPS-CMs developed significant prolongation in FPDc after treatment with 0.1 and 1 μM of E4031, 10 μM of vandetanib, and 100 μM of clarithromycin (Figure 6A-C). These results show that the particular susceptibility to develop abnormal repolarization in response to sotalol extends to other hERG blockers. Cardiomyocytes from patients presenting high sensitivity to sotalol exhibit similar sensitivity to other hERG blockers. One may speculate whether the abnormal response to these hERG blockers is also mediated by CAVIN1. In line with the observations reported with sotalol, CAVIN1 overexpression in LS iPS-CMs or silencing in HS iPS-CMs mediated dramatic changes in the response to these hERG blockers. CAVIN1 overexpression induced a more significant change in the cardiac repolarization duration of HS iPS-CMs than of LS iPS-CMs after treatment with three hERG blockers (Figure 6D-F). Reciprocally, CAVIN1 silencing abrogated the reactivity to increasing concentrations of hERG blockers in HS iPS-CMs (Figure 6G-I). Taken together, cardiomyocytes presenting high sensitivity to sotalol are also sensitive to other hERG blockers, and CAVIN1 expression level drives the cellular responses to these blockers.

**Figure 6:**
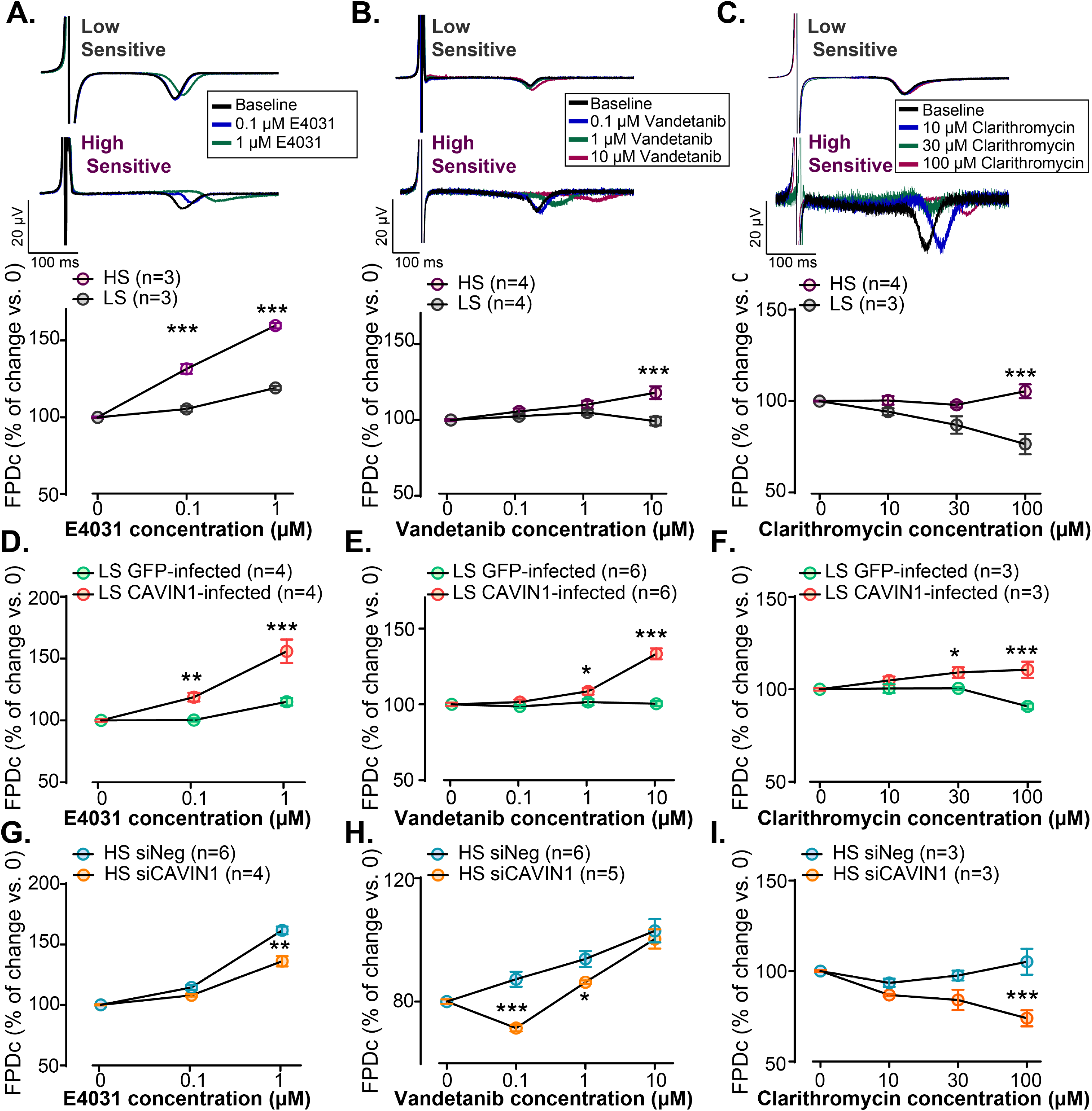
Universal mechanism implicating CAVIN1 in response to other hERG blockers. A. Top: Representative superimposed non-corrected FP recorded from LS and HS iPS-CMs following treatment with different concentrations of E4031. Bottom: Averaged percentage of change in FPDc (corrected to beating frequency) as compared to that at baseline following application of 0.1 and 1 μM E4031. *** p < 0.001 *vs*. LS iPS-CMs (two-way ANOVA with Bonferroni post-hoc test). B. Top: Aligned non-corrected FP recorded from LS and HS iPS-CMs treated with different concentrations of vandetanib. Bottom: Averaged percentage of change in FPDc (corrected to beating frequency) as compared to that at baseline following application of 0.1, 1, and 10 μM of vandetanib. *** p < 0.001 *vs*. LS iPS-CMs (two-way ANOVA with Bonferroni post-hoc test). C. Top: Aligned non-corrected FP recorded from LS and HS iPS-CM treated with different concentrations of the antibiotic, clarithromycin. Bottom: Averaged percentage of change in FPDc (corrected to beating frequency) as compared to that at baseline following application of 10, 30, and 100 μM of clarithromycin. *** p < 0.001 *vs*. LS iPS-CMs (two-way ANOVA with Bonferroni post-hoc test). D, E, and F Averaged percentage of change in FPDc as compared to that at baseline measured in LS iPS-CMs infected with either GFP- or CAVIN1-T2A-GFP adenoviruses in response to increasing concentrations of E4031, vandetanib, and clarithromycin. *, **, and *** p < 0.05, p < 0.01, and p < 0.001 *vs*. GFP-infected LS iPS-CMs, respectively (two-way ANOVA with Bonferroni post-hoc test). G, H, and I Percentage change in FPDc as compared to that at baseline following application of different concentrations of (from left to right) E4031, vandetanib, and clarithromycin at day 5 of transfection of HS iPS-CMs with either siNeg- or CAVIN1-siRNA. *, **, and *** p < 0.05, p < 0.01, and p < 0.001 *vs*. GFP-infected LS iPS-CMs, respectively (two-way ANOVA with Bonferroni post-hoc test).

## DISCUSSION

The present study uncovers a new role of CAVIN1 in mediating susceptibility to diLQT syndrome. Higher expression of CAVIN1 promoted higher turnover of hERG channels from the membrane to the cytoskeleton in response to hERG blockers, consequently leading to an excessive prolongation of the repolarization phase. CAVIN1 expression was significantly higher in cardiomyocytes derived from subjects predisposed to diLQT *in vivo*, and CAVIN1 expression modulation was sufficient to shift the cell’s response to hERG blockers. A progressive prolongation of the repolarization phase was observed in LS iPS-CMs with increasing Sotalol concentrations, likely reflecting the direct pharmacological inhibition of hERG. In contrast, the higher response to low-concentrations of Sotalol observed in HS iPSC-CMs resulted from an additional mechanism associated with an unbridled internalization of hERG via a CAVIN1/caveolae-dependent process. Variants in *CAVIN1* cause lipodystrophy and have been previously associated with congenital LQT syndrome, but the precise mechanism is yet questionable^16^. CAVIN1 exerts a pivotal role in the biogenesis of caveolae, which are membrane invaginations enriched in cholesterol^17^ and expressing hERG channels^18, 19^. CAVIN1 expression level is positively correlated with the number of caveolae at the cardiomyocytes’ membrane ^20^, consistent with our results showing elevated expression of CAV1 and CAV3, the signature proteins of caveolae, in CAVIN1-infected cells and reduction in caveolae density at the membrane after CAVIN1 knockdown. Intriguingly, *CAV3* variants have been associated with congenital long QT syndrome type 9 and CAV 3 can affect the availability of hERG channels at the cell membrane^21^. Previous studies have reported that the clathrin-independent pathway, including caveolae, mediates hERG endocytosis under physiological conditions^19, 22-25^. Our results demonstrate the involvement of caveolae- and CAVIN1-dependent endocytic route of hERG channels in response to hERG blocker exposure. This endocytic route is rapidly activated in the presence of the blocker and is pathologically excessive in high-sensitive individuals that are thus predisposed to develop diLQT.

The dynamics of caveolae are regulated by the elements of the cytoskeleton, notably microtubules and actin filaments. CAV1 explicitly mediates the interaction between caveolae and cytoskeleton and is, therefore, involved in vesicular trafficking ^26-28^. We observed expression of caveolae markers in the cytoskeleton protein fraction of iPS-CMs, suggesting the direct communication between the membrane and the cytoskeleton as two key endocytic compartments for hERG channels. Inhibition of caveolae endocytosis was able to decrease to excessive repolarization response to Sotalol. Our results suggest that CAVIN1 promotes a rapid transfer of hERG channels from the plasma membrane to the cytoskeleton after sotalol application. Once internalized, membrane proteins undergo different steps, including trafficking to degradation or recycling back to the membrane surface ^29^. We observed an accumulation of the cleaved form of hERG (70 kDa) ^30-32^ in HS iPS-CMs after sotalol exposure, suggesting that hERG channels are, at least in part, degraded after CAVIN1-dependent endocytosis. *CAVIN1* silencing was associated with a reduction in fragmented hERG level. The synthesis process of HERG channels is slow, and hERG recycling back to the membrane is essential to maintain its stable surface expression and ensure fast recovery of the repolarization duration ^33^. Trafficking toward the membrane is notably mediated by the GTPase RAB11 ^33-36^. Our protein fractioning data showed higher recruitment of RAB11 to the membrane in HS iPS-CMs than in LS iPS-CMs after treatment with sotalol. These results are indicative of an increase in recycling endosomes and acceleration of the protein turnover to rescue hERG density at the surface membrane, and may explain why QT interval returns to normal after discontinuation of the drug.

Previous studies have attempted to improve our understanding of the genetic predisposition to diLQT ^6, 8, 9^. Despite some association with genetic variants, especially on the *KCNH2* gene, the vast majority of diLQT cases remain genetically unresolved ^8^. In contrast to these studies, we herein identified higher expression of CAVIN1 as a key molecular mechanism to explain the repolarization delay in response to hERG blockers. Hence, future studies should be devoted to understanding the regulation of CAVIN1 expression and why CAVIN1 expression is physiologically higher in some individuals. There are currently no known genetic variants of *CAVIN1* associated with the regulation of its expression. Moreover, the impact of sex hormones on CAVIN1 expression is undetermined and deserves further investigation. It is indeed well established that women are at a higher risk of diLQT than men and that contraceptive pills can aggravate this risk ^37^. Consistently, female sex hormones such as progesterone were shown to alter hERG distribution by disrupting the homeostasis of cholesterol—the main constituent of caveolae ^38^. Whether this process depends upon CAVIN1 remains to be determined.

As per the ICH S7B/E14 guidelines, evaluation of the potential of a novel drug to delay cardiac repolarization is a crucial step in drug development. In this regard, the ability to reduce I_Kr_ is specifically addressed in animal and *in vitro* models, typically heterologous expression systems of ion channels ^39^ which, however, are limited by their capacity to recapitulate the complex human cardiac electrophysiology and the interindividual variability to develop cardiotoxic side-effects ^10, 11, 40, 41^. Consequently, many drugs have escaped preclinical elimination and therefore disposed consumers to fatal arrhythmias; some have been ultimately withdrawn from the market ^42, 43^. Recently, the Comprehensive *in vitro* Proarrhythmia Assay (CiPA) added hiPS-CMs to drug safety evaluation ^44^, as they provide a more reliable representation of human cardiomyocytes ^44^. In line with our previous results ^10^, here we show that the cardiotoxic response of iPS-derived cells is variable and depends on the predilection to diLQT, as observed in the subject from whom iPSCs were generated. This panel further served to identify an original molecular mechanism that supports the inter-subject variability in cardiotoxic response to hERG blockers. In addition, this hiPSC platform offers the unique opportunity to test the response to multiple offending drugs, which will not be possible in patients with proven diLQT. We critically observed for the first time a cross-sensitivity to other drugs targeting hERG (such as vandetanib, E4031, and clarithromycin) that was also dependent on CAVIN1 expression. This observation also suggests that the activation of the CAVIN1 system is likely triggered when the hERG channel activity is reduced rather than by a specific binding of drugs to the channel, but this would deserve further investigations. Altogether, these findings emphasize the potential of using patient-specific iPS-CMs for drug safety screening, modeling of inter-individual variability, and identification of molecular culprits.

Our study has some limitations that should be acknowledged. The level of CAVIN1 expression within iPS-derived cardiomyocytes appeared as a critical marker of the susceptibility to develop diLQT. At present, there is no accurate way to estimate CAVIN1 expression in the human myocardium. Additional investigations will be warranted to establish the translational value of prior estimation of CAVIN1 expression in candidates to receive a hERG blocker. The acceleration of the channel degradation was described in response to other stimuli such as hypokalemia ^22, 45, 46^ or drugs ^19, 47^; whether CAVIN1 controls these processes is yet to be established.

In conclusion, this study identified a common molecular modulator of the susceptibility to diLQT in human iPS-CMs derived from individuals with different genetic backgrounds. CAVIN1 controlled membranous hERG turnover upon pharmaceutical stimulation, and therefore the repolarization duration. Different expression levels of this marker may explain the inter-individual variability in response to hERG blockers. Thus, these results classify CAVIN1 as a potential risk factor for diLQT and open perspectives for a personalized drug prescription to avoid undesirable cardiotoxicity.

## Data Availability

The data and material that support the findings of this study are available from the corresponding author upon reasonable request.

